# Recurrent aphthous stomatitis affects quality of life

**DOI:** 10.1101/2022.04.01.22273300

**Authors:** César Rivera, Mariagrazia Muñoz-Pastén, Esteban Muñoz-Núñez, Romina Hernández-Olivos

## Abstract

Recurrent aphthous stomatitis are recurrent oral ulcers that can affect important daily activities, such as oral hygiene and eating. In this prospective case-control study (n=62), we show that, during ulcer episodes, patients report a poorer quality of life compared to ulcer-free periods, and that this impact is positively associated with the number and size of lesions. Our results suggest that, if intervened locally, general relief of the condition could be achieved.

## INTRODUCTION

Recurrent aphthous stomatitis (RAS) is the most common disease of the oral mucosa. It is characterized by recurrent, painful, single or multiple ulcers, with erythematous margins (1). The disease sequence comprises several stages, including premonitory, pre-ulcerative, ulcerative, healing, and remission phases (2). The ulcerative phase (presence of active lesions) and remission (without evidence of lesions) are the stages that can be evaluated more objectively in the clinical examination (1).

RAS can interfere with important functions such as talking, eating, swallowing, and consequently affect quality of life (3). Quality of life is a concept that refers to the general well-being of an individual, including physical, emotional, and psychological parameters. This concept is often used to describe the impact of a disease or the effects of a medical intervention on general health (4).

Despite being a frequent disease, the low number of studies available evaluating the impact of ulcers on quality of life is surprising. In fact, there are only two investigations that evaluate this impact of disease activity, that is, when the lesions are present and then in an ulcer-free period (3, 5). However, it is not known whether the local clinical characteristics of the lesions (eg number and size of lesions) could explain the observed impact levels. Therefore, the objective of this research is to determine the impact of RAS on quality of life related to oral health, and secondly to determine the correlation between the observed impact and lesions characteristics.

## METHODS

### General design

Our investigation corresponded to a case-control study. In it we evaluate the clinical characteristics of RAS and their impact on quality of life. For this, we record the quality of life reported in healthy people (without a history of ulcers) and people with RAS. Furthermore, in the latter we recorded the quality of life during the disappearance phase of the lesions. Finally, we correlate the clinical characteristics of the lesions with the levels of quality of life. All procedures followed the guidelines of the Declaration of Helsinki (6) and Ethics committee/IRB of University of Antofagasta gave ethical approval for this work (folio 156/2018, https://doi.org/10.6084/m9.figshare.11225900).

### Patients

We prospectively evaluated 62 patients in our clinical service (Center of Dental Clinics of the University of Talca) between March and October 2019. The patients were divided into a “healthy” group (n = 29, people with no history of ulcers) and “RAS” (N = 33, people who had an active ulcer at study entry). The latter were also evaluated when the lesion disappeared (n = 33, remission). We conducted a total of 95 clinical examinations.

### Inclusion and exclusion criteria

To be included, the patients had to present ulcerative lesions with a history of no more than 3 days (7). Furthermore, these lesions must have had an anatomical location that allowed them to be photographed. Exclusion criteria were the use of medications to treat ulcers and the use of topical or systemic corticosteroids. Other exclusion criteria were the presence of other types of lesions of the oral mucosa, diseases that present with acute or chronic pain, smoking, excessive alcohol consumption (more than three times a week) (8) and the presence of any of the following conditions at the time of the examination: Behçet syndrome, folic acid deficiency, iron deficiency anemia, vitamin B-12 deficiency, Crohn’s disease, ulcerative colitis, lichen planus, pemphigus, pemphigoid, oral herpes, herpangina, iron deficiency, systemic lupus erythematosus, reactive arthritis, oral candidiasis, celiac disease, cyclic neutropenia, gluten-sensitive enteropathy, malabsorption, pernicious anemia, Sweet’s syndrome (acute febrile neutrophilic dermatosis), Marshall syndrome, or PFAPA syndrome (periodic fever, aphthous stomatitis, pharyngitis and adenitis), trisomy 8, severe periodontal disease, erythema multiforme, syphilis and HIV-AIDS. For inclusion as controls, patients should never have had thrush. Except for the absence of ulcers, this group shared the same inclusion and exclusion criteria.

### Episode characteristics

Diagnosis was established by a clinical examination by a doctor in oral medicine. We classify the lesions according to the standard pathological nomenclature (minor, major or herpetiform ulcer) (9-12). In addition, we asked patients how many episodes of ulcers they suffered in the last year. We recorded the number of lesions present on examination, total ulcerated diameter, mean diameter of lesions, and pain associated with ulcers (using a visual analog scale, VAS).

### Quality of life

We determined the impact of the disease on the quality of life related to oral health in healthy controls and patients with thrush (during the injury and its remission) by applying the impact profile of oral health in its abbreviated Spanish version (OHIP-14SP) (13). OHIP-14SP explores seven dimensions: functional limitation, physical pain, psychological discomfort, physical disability, psychological disability, social disability and handicap. They are evaluated through questions of 5 levels (never, almost never, occasionally, frequently and very frequently) with a score from 0 to 4. The total OHIP score is between the ranges of 0 and 56 points (sum of all the answers). To view the OHIP-14SP questionnaire, see Supplementary Table S1 (https://doi.org/10.6084/m9.figshare.11225900).

### Statistical analysis

Results were presented as frequencies and means ± standard deviation. We analyzed the differences between the groups using one-way ANOVA with post hoc Tukey and Games-Howell contrasts. To analyze the association between quantitative variables we used Spearman’s correlation coefficient. In all procedures we use a confidence level of 95% (p-value ≤ 0.05).

## RESULTS

### Quality of life

We included 62 patients in our research. They were divided into two groups: healthy (n = 29) and RAS (n = 33). The mean age was 26 years for both, with women being the majority (24 and 27 respectively). All the lesions were of the minor morphological type. Most of the lesions were located on the lining mucosa, preferably on the lips. Table I shows clinicopathological characteristics associated with presence of ulcers. The RAS group was in turn divided into lesion (active ulcer presented at study entry) and remission (when the lesion disappeared completely). All the participants answered the quality of life questionnaire. The score was noticeably higher when there were lesions (Figure 1).

**Table I.** Characteristics of recurrent aphthous stomatitis episodes.

| Variable | Magnitude |
| --- | --- |
| Ulcer episodes in the past year | 7.1±5.8 |
| Number of lesions on examination (n) | 1.4±0.7 |
| Total ulcerated diameter (mm) | 3.8±2.0 |
| Mean diameter (mm) | 3.3±1.4 |
| Reported pain (EVA) | 4.2±2.1 |

Numbers represent means ± standard deviation.
Subjects without RAS reported no pain

**Figure 1.**
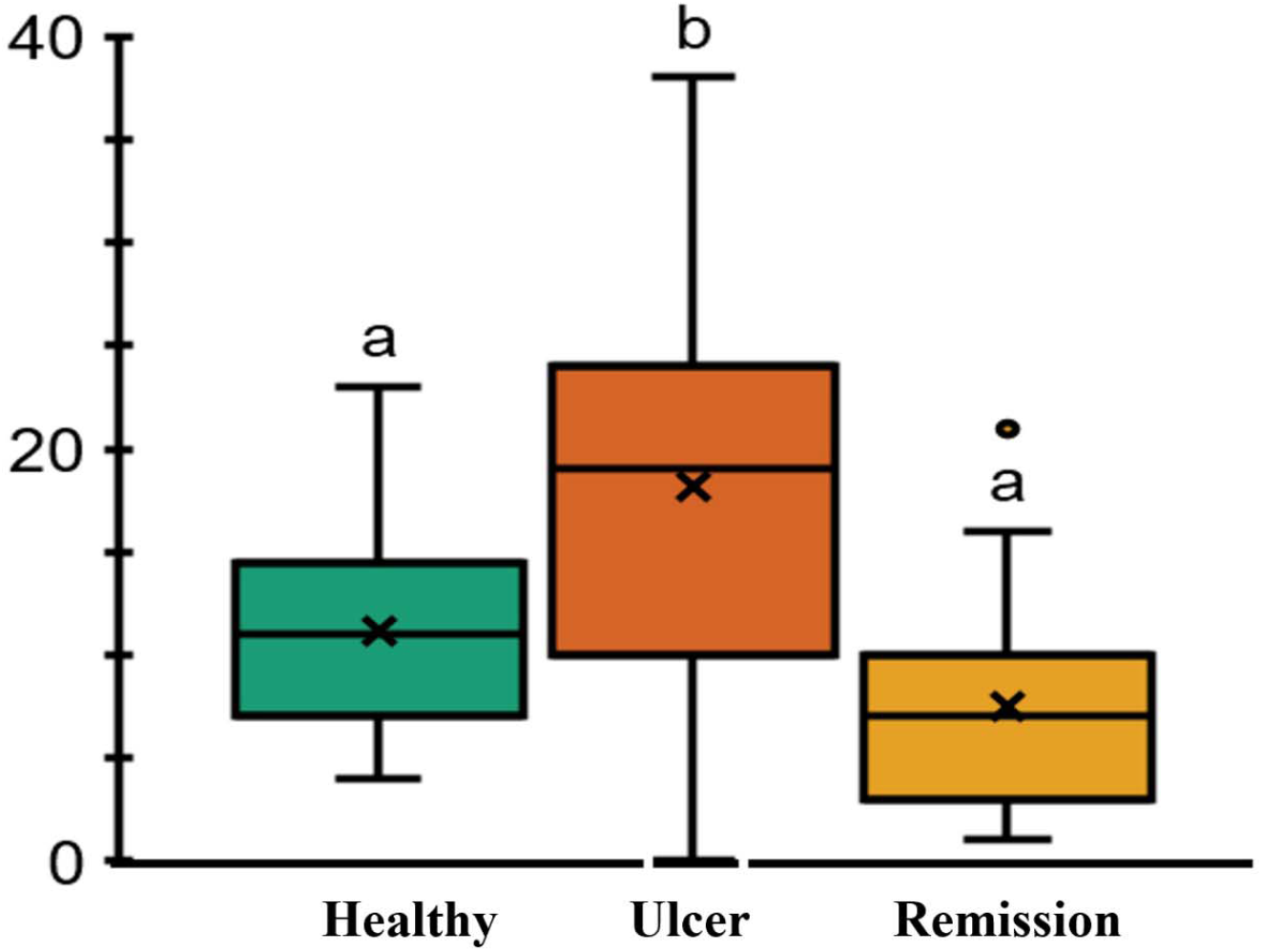
Recurrent aphthous stomatitis affects quality of life. Total OHIP-14Sp score. On the box-and-whisker plot, the boxes represent quartile 1, median, and quartile 3. The x stands for the mean. The whiskers extend to the maximum and minimum values. Data outside of these limits represents outliers. The different letters indicate a statistically significant difference (p-value <0.05, one-way ANOVA with Tukey’s post hoc contrasts).

### Distribution of OHIP-14Sp in the presence and absence of oral ulcers

Table II illustrates the distribution of OHIP-14Sp scores in the healthy and RAS groups. In general, the presence of lesions presents higher means when compared with subjects in healthy and in remission. It is striking that healthy patients have greater difficulty resting, which can be explained because they were mainly university students.

**Table II.**
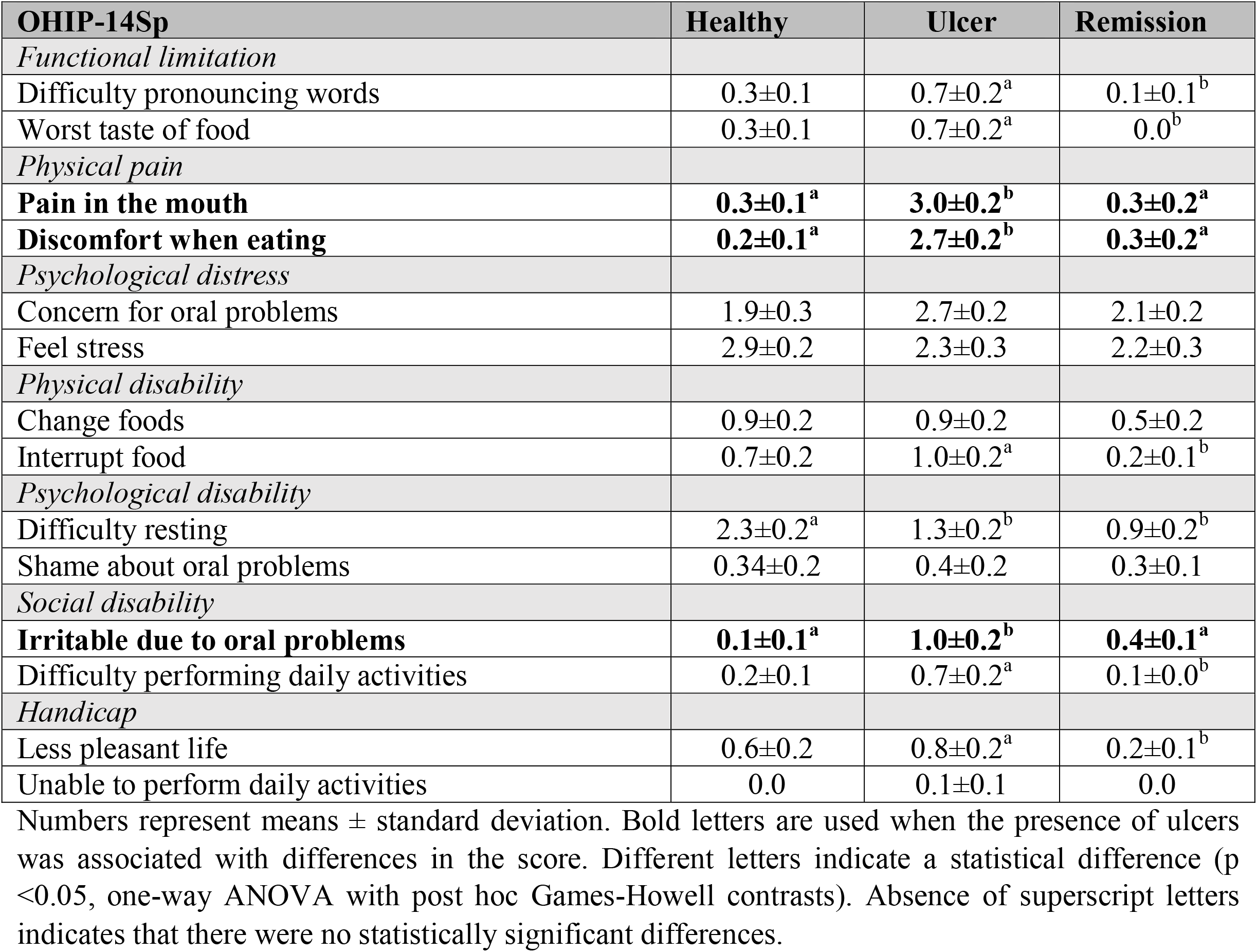
Scores of the OHIP-14SP items.

### OHIP-14Sp dimensions and lesion characteristics

Table III shows the relationship between the OHIP-14Sp items altered by the presence of RAS and the clinicopathological variables evaluated in the ulcers. The mean size of the lesions in the oral mucosa and the number of ulcers are the clinical variables that are best related to pain in the mouth, discomfort when eating and irritability.

**Table III.** Number and size of recurrent aphthous stomatitis lesions affect quality of life.

| OHIP-14Sp | Spearman's correlation coefficient |  |  |
| --- | --- | --- | --- |
|  | Number of ulcers | Ulcerated total diameter | Mean size |
| Mouth pain | 0.8** | 0.5** | 0.8** |
| Discomfort when eating | 0.7** | 0.6** | 0.8** |
| Irritable due to oral problems | 0.4** | 0.4* | 0.5** |

Degree of relationship between dimensions affected by recurrent aphthous stomatitis and clinical characteristics of lesions. A coefficient equal to or greater than 0.8 indicates a good correlation. \*\* Correlation is significant at p-value <0.01 and \*p-value <0.05.

## DISCUSSION

Quality of life assessments have an important place in health care. In recent years, they have become an accepted end point in clinical research trials (5). Individual perceptions of the impact profile on oral health have grown in importance and may have a direct effect on people’s health-related quality of life (14). We are interested in evaluating the impact of RAS on the quality of life of those who suffer from it, to establish firstly whether the impact depends on the activity of the disease and secondly whether this effect may be associated with some local characteristic of injuries. In our research, we observed that RAS negatively affect quality of life, causing pain mainly associated with the number of ulcers and the average size of lesions.

Patients reported presenting more than 7 episodes of ulcers per year. Considering that these lesions resolve in an approximate period of 2 weeks, a participant may be more than 27% of a current year with the discomforts that are associated with the disease. This emphasizes the need to pay attention to how ulcers compromise well-being.

Our sample is representative of the reality reported for this disease in the world. All of our participants presented minor lesions located on non-keratinized mucosa, mainly on the lips. This is the most frequent and least severe ulcer form (14). They are generally located on the lips, tongue, and oral vestibule (15). The fact that our patients correspond to the classically described pattern allows us to trust that the conclusions obtained from our data can be extrapolated to other populations.

RAS affected the quality of life of patients. To evaluate the effect of the lesions, we used the OHIP-14 scale. This is the most widely used instrument to assess the impact of oral health on quality of life (16). Along with examining healthy subjects as a basis, we applied the questionnaire in two disease periods, when lesions were present and then in the remission phase. Measuring the quality of life in different states of ulcer activity in the same subjects allowed us to have a better control of the design, since the changing condition is the presence or absence of the lesions. This provides a correct and isolated measure of the effects of ulcers (3).

Notoriously, presence of RAS obtained much higher OHIP-14 scores, compared to healthy subjects and subjects whose ulcers disappeared. Five articles have previously evaluated the impact of RAS on quality of life, all finding a negative effect (3, 5, 17-19). Of these, 2 performed it in active lesions and in ulcer-free periods (3, 5). Like us, they observed that the quality of life associated with oral health when ulcers are present is significantly lower. In this way, we went further in trying to understand what the impact is due to. Our findings showed that RAS caused pain, discomfort when eating, and irritability. This was positively associated with the number of ulcers present in the oral mucosa and average size of lesions. The impact was greater, while the number of ulcers and their size was also. This result shows that, if the condition caused by oral ulcers is to be improved, therapeutic measures must be focused on treating the local compromise of oral mucosa. Unfortunately, the evidence available today for the successful management of lesions is insufficient, both for topical and systemic treatments (20, 21).

Our results are limited to the fact that the impact of the lesions was essentially measured with self-reports obtained from an instrument that was designed for dental diseases and not for oral mucosa. New methods are being developed to evaluate the affection produced by mucosal lesions (22), however, its extension has not reached the use of OHIP-14.

In this research, we concluded that RAS increased the negative effects of oral health on the quality of life of patients. The number and size of the injuries are responsible for this impact. Intervening on these local characteristics could alleviate pain, eating difficulties, and irritability in affected patients.

## Data Availability

All data produced in the present work are contained in the manuscript.

## Acknowledgements

We want to thank all the volunteers who actively participated in this research, despite the national social context and the global pandemic, and the University Dental Clinic Center for hosting us. We also want to give special recognition to the TENS Yennifer Lemus.

## Funding

This research was funded by the Chilean National Agency for Research and Development (ANID) FONDECYT Iniciación grant number #11180170 (to C.R.) and Concurso de Proyectos de Investigación de Alto Nivel en Odontología, Red Estatal de Odontología grant number #REO19–012 (to C.R.).

## Disclosure

All the authors declare that they have no competing interests.

## Author contribution statement

R.H., M.M., E.N., E.A and C.R. contributed to data acquisition and critically revised the manuscript. C.R. contributed to conception, design, project supervision, performed all statistical analyses, and interpretation, drafted and critically revised the manuscript. All authors gave their final approval and agreed to be accountable for all aspects of the work.

## Notes

### Competing Interest Statement

The authors have declared no competing interest.

### Funding Statement

This research was funded by the Chilean National Agency for Research and Development (ANID) FONDECYT Iniciacion grant number #11180170 (to C.R.) and Concurso de Proyectos de Investigacion de Alto Nivel en Odontologia, Red Estatal de Odontologia grant number #REO19-012 (to C.R.).

### Author Declarations

Ethics committee/IRB of University of Antofagasta gave ethical approval for this work (folio 156/2018, https://doi.org/10.6084/m9.figshare.11225900).

